# Integrated genomic analysis and CRISPRi implicates *EGFR* in Alzheimer’s disease risk

**DOI:** 10.1101/2025.06.25.25328705

**Authors:** Yuk Yee Leung, Pavel P. Kuksa, Luke Carter, Jeffrey Cifello, Emily Greenfest-Allen, Otto Valladares, Louisa Boateng, Shannon Laub, Natalia Tulina, Sofia Moura, Aura Ramirez, Katrina Celis, Fulai Jin, Ru Feng, Gao Wang, Phil De Jager, Jeffery M Vance, Liyong Wang, Struan F.A. Grant, Gerard D Schellenberg, Alessandra Chesi, Li-San Wang

## Abstract

Genome-wide association studies (GWAS) have identified numerous loci linked to late-onset Alzheimer’s disease (LOAD), but the pan-brain regional effects of these loci remain largely uncharacterized. To address this, we systematically analyzed all LOAD-associated regions reported by Bellenguez et al. using the FILER functional genomics catalog across 174 datasets, including enhancers, transcription factors, and quantitative trait loci. We identified 42 candidate causal variant-effector gene pairs and assessed their impact using enhancer-promoter interaction data, variant annotations, and brain cell-type-specific gene expression. Notably, the LOAD risk allele of rs74504435 at the *SEC61G* locus was computationally predicted to increase *EGFR* expression in LOAD related cell types: microglia, astrocytes, and neurons. Functional validation using promoter-focused Capture C, ATAC-seq, and CRISPR interference in the HMC3 human microglia cell line confirmed this regulatory relationship. Our findings reveal a microglial enhancer regulating *EGFR* in LOAD, suggesting *EGFR* inhibitors as a potential therapeutic avenue for the disease.

## BACKGROUND

Alzheimer’s disease (AD) is the leading cause of dementia in the United States and currently lacks effective treatments or prevention strategies. The most common form, late-onset Alzheimer’s disease (LOAD), typically begins after age 60 and is highly heritable (60–80%), indicating a significant genetic component in its development^1^. While the *APOE* locus remains the strongest genetic risk factor^2^, LOAD is complex and highly polygenic^3^. Previous genome-wide association studies (GWAS) identified over 20 LOAD-associated loci^2,4^; recent studies using UK Biobank proxy-AD or proxy-control samples have expanded this list to 75 loci^5,6^. Although progress has been made in linking LOAD genetic risk to microglial-mediated innate immune processes^7–9^, the broader cellular contexts of these variants remain incompletely understood. Emerging evidence suggests that LOAD-associated variants also affect other brain cell types, including myeloid cells^10^, astrocytes and neurons, but the mechanisms across these diverse cellular environments remain largely uncharacterized.

Over 90% of GWAS variants are located in non-coding regions of the genome, outside of protein-coding sequences^10,11^. These non-coding variants are widely hypothesized to affect gene regulatory elements, such as enhancers^12–14^, which can influence the expression of distant target genes^15^. The difficulty in identifying such distal genes arises from the challenges related to linkage disequilibrium (LD) with nearby non-causal variants and the variability in the biological contexts of the corresponding target ‘effector’ genes. Despite these, some studies have applied various statistical and computational methods, along with new data types, to analyze non-coding GWAS signals for AD^10,16–18^. This effort is important because drugs with genetic support are twice as likely to gain approval^19–21^.

However, these studies face limitations in their analytical strategies. First, they traditionally focus only on top GWAS signals (sentinel variants), restricting the understanding of AD’s full genetic landscape^5,6,16^. Since any variant in LD with a sentinel could be causal, comprehensive analyses should include both sentinel and nearby LD variants. Second, genome-wide functional genomics data for brain tissues or specific cell types remain limited and relatively small compared to data from other cell types or cell lines ^22–24^. Third, prior computational analyses of non-coding variants often failed to integrate eQTLs from independent sources to confirm consistency and replication of effect alleles and signal direction ^5,6,10,11^. Finally, no previous analyses have combined eQTLs and enhancer-promoter interactions (EPI) to prioritize variant-to-gene (V2G) pairs in LOAD.

To address these limitations, we developed an enhanced post-GWAS non-coding variant analysis framework^25^. This approach systematically identifies candidate causal variants and relevant regulatory genomic features to improve our understanding of genetic loci associated with LOAD.

## RESULTS

We summarize our strategy in **Fig. 1**, which comprises four key steps: “Preprocessing,” “Characterization,” “*In silico* validation,” and “Functional validation”.

1) In the “Preprocessing**”** stage, we obtained pairwise-independent tag variants through LD-based pruning (1000 Genome panel) on all genome-wide significant LOAD GWAS variants (*p*<5×10^-8^)^5^. To produce a larger pool of candidate causal variants, we included all proxy variants through LD-based expansion of the tag variant set (r^2^>=0.7).
2) In the “Characterization” step, these variants were annotated using FILER^26^ (a large-scale genomic data queryr tool), and their potential functional context(s) were predicted using SparkINFERNO^25^ (a scalable pipeline for inferring non-coding variants molecular mechanisms). We identified candidate causal genes per variant using eQTL data. Additionally, we used HOMER^27^ to predict transcription factor binding site (TFBS) disruptions caused by the variants. Only variants within brain enhancers were retained in subsequent analysis steps. Together with their corresponding genes, this set formed the enhancer-based causal V2G pairs with TFBS.
3) For the “*In silico* validation” step, we annotated and ranked the V2G pairs, putative causal variants, and effector genes using independent assays from new data sources. These include regulatory features (EPIs, QTLs, and open chromatin regions) and expression datasets (bulk RNA-seq, proteomics). We also performed consistency checks on QTLs across data sources.
4) Finally, in the “Functional validation” step, we specifically contextualized one V2G pair using our existing datasets from multiple cell types, and *in vitro* validation using CRISPR interference (CRISPRi).

**Fig. 1:**
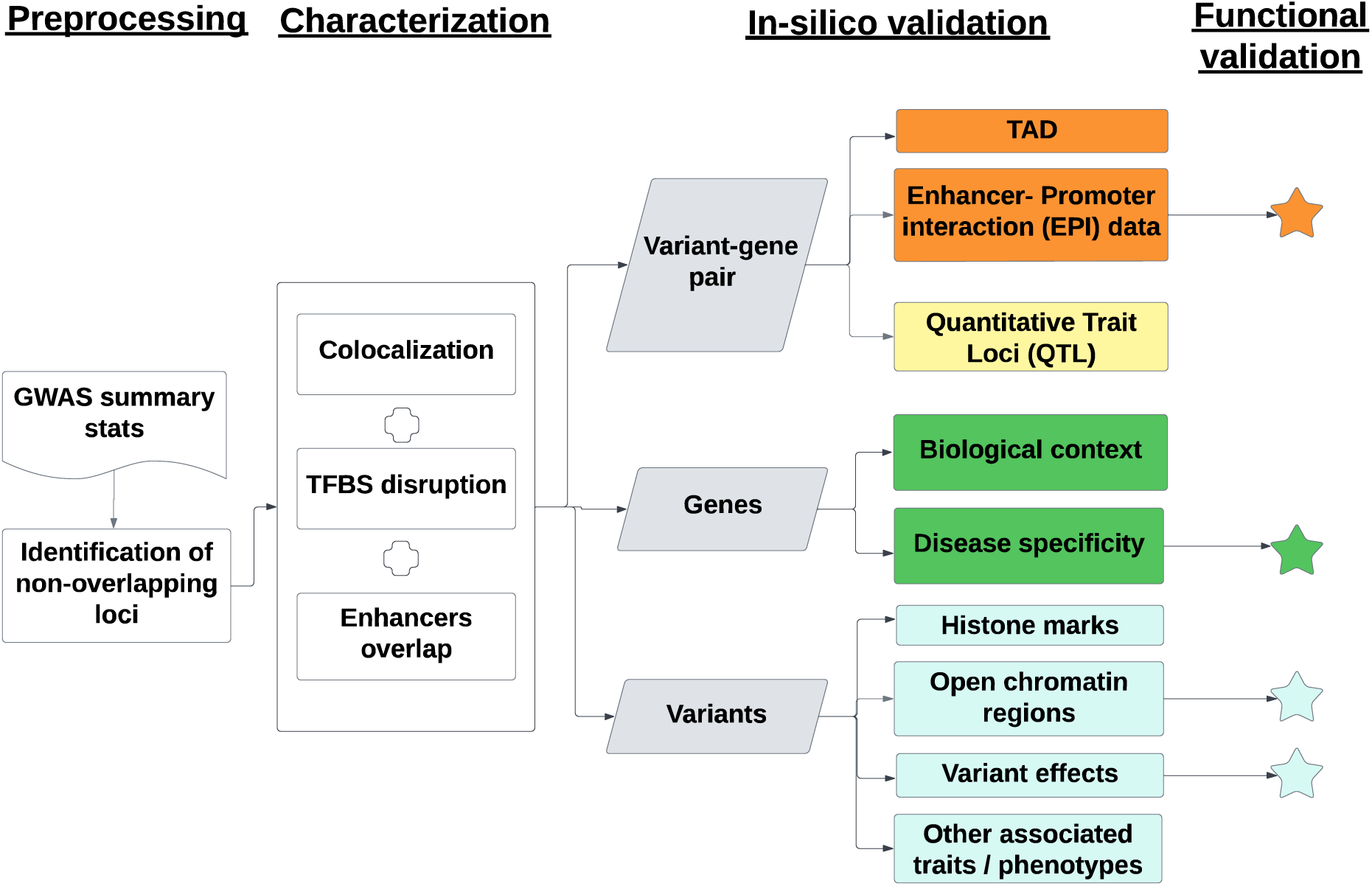
Analysis strategy. Our post-GWAS framework consists of the following steps: “Preprocessing,” “Characterization,” “In silico validation,” and “Functional validation.” Genome-wide significant variants from Bellenguez *et al.* 2022 were leveraged to identify regions of interest. Putative causal variants were analyzed for functional contexts and linked to potential causal genes using enhancers, eQTL data and TFBS predictions. These V2G pairs were characterized in brain tissues and cell types. Independent assays including enhancer-promoter interactions, QTLs, and open chromatin regions from new data sources were used for *in silico* validation. Functional validation included promoter-focused Capture C, ATAC-seq, RNA-seq, and CRISPRi in microglial cells.

Overall, we defined context-specific regulatory elements (variants and genes) across different cellular and tissue contexts leveraging 174 datasets from 10 data sources. This enabled independent *in silico* validation of 42 variant-effector-gene (V2G) pairs. The rs74504435-*EGFR* V2G pair underwent further functional characterization, confirming it as a therapeutically tractable target.

### Preprocessing: identification of genomic regions and candidate variants of interest

To define regions of interest and establish an initial discovery set of plausible candidate variants for further analyses, we leveraged the set of 5,586 genome-wide significant variants (*p*<5×10^-8^) from the full GWAS summary statistics^5^. We performed LD pruning of using the 1000 genomes EUR panel^28^, obtaining 580 pairwise-independent (tag) variants (r²<0.7). For each tag variant, we expanded its region of interest to include all proxy variants (r² ≥ 0.7) within 1 Mbps, with boundaries set by the most distant proxies. This LD-based expansion yielded a total of 9,762 plausible candidate variants across all identified regions of interest (*Preprocessing in **Fig. 1**, Methods: Preprocessing*), increasing the candidate pool by 75% (from 5,586 to 9,762). Most candidate variants were non-coding, predominantly intergenic (38%) or intronic (24%), with 35% located in 5’ and 3’ UTR introns (**Supplementary Fig. 1**).

### Characterization: identification of putative causal variants, genes, and variant-gene pairs

To explore the regulatory roles of candidate variants, we adapted the SparkINFERNO framework^25^ and overlapped variants with 174 brain-related functional genomics (FG) tracks in FILER^26^. These tracks cover 35 brain regions and 7 brain cell types across 10 data resources, representing five regulatory types: enhancers, histone modifications, QTLs, EPIs and topologically associating domains (TADs) (**Supplementary Fig. 2**).

We quantified the probability of a candidate regulatory variant co-localizing with an eQTL signal. Among 9,762 variants (including 130 on chr19 in the APOE region), 393 had at least one colocalized signal (coloc_PP.H4.abf > 0.7) in any of 13 GTEx^29^ brain tissue-specific eQTL tracks with nominally significant results. These 393 variants defined 2,763 tissue-V2G pairs (comprising 229 independent tag variants), putatively regulating 338 genes. Notably, 25 variants were identified in ≥10 brain regions, while 182 variants (46%) were brain-region specific. **Fig. 2A** highlights the top colocalization results (defined by locus-level posterior colocalization probability > 0.99 and fewer than 100 SNPs per locus) across GTEx brain tissues.

**Fig. 2.**
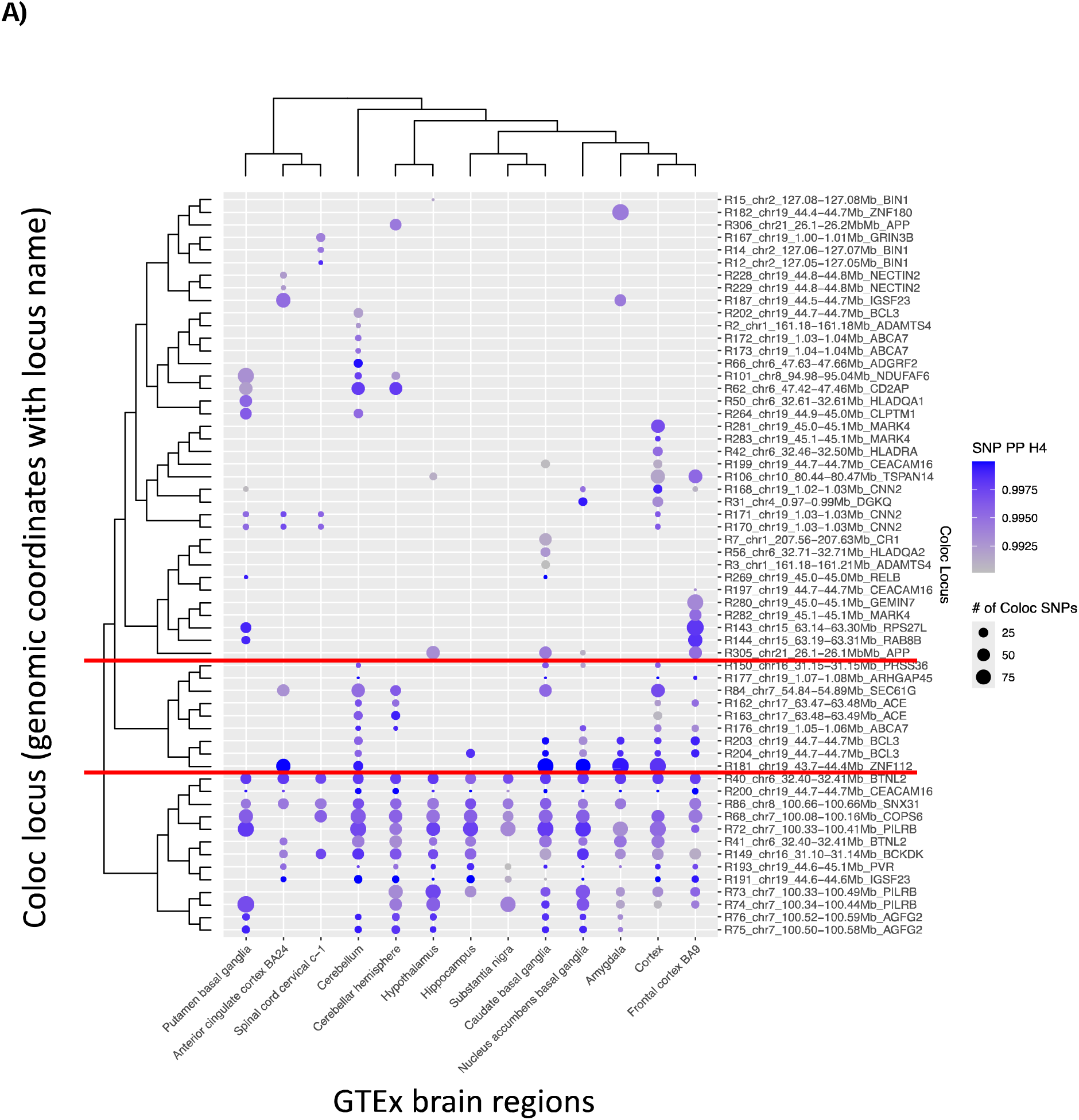
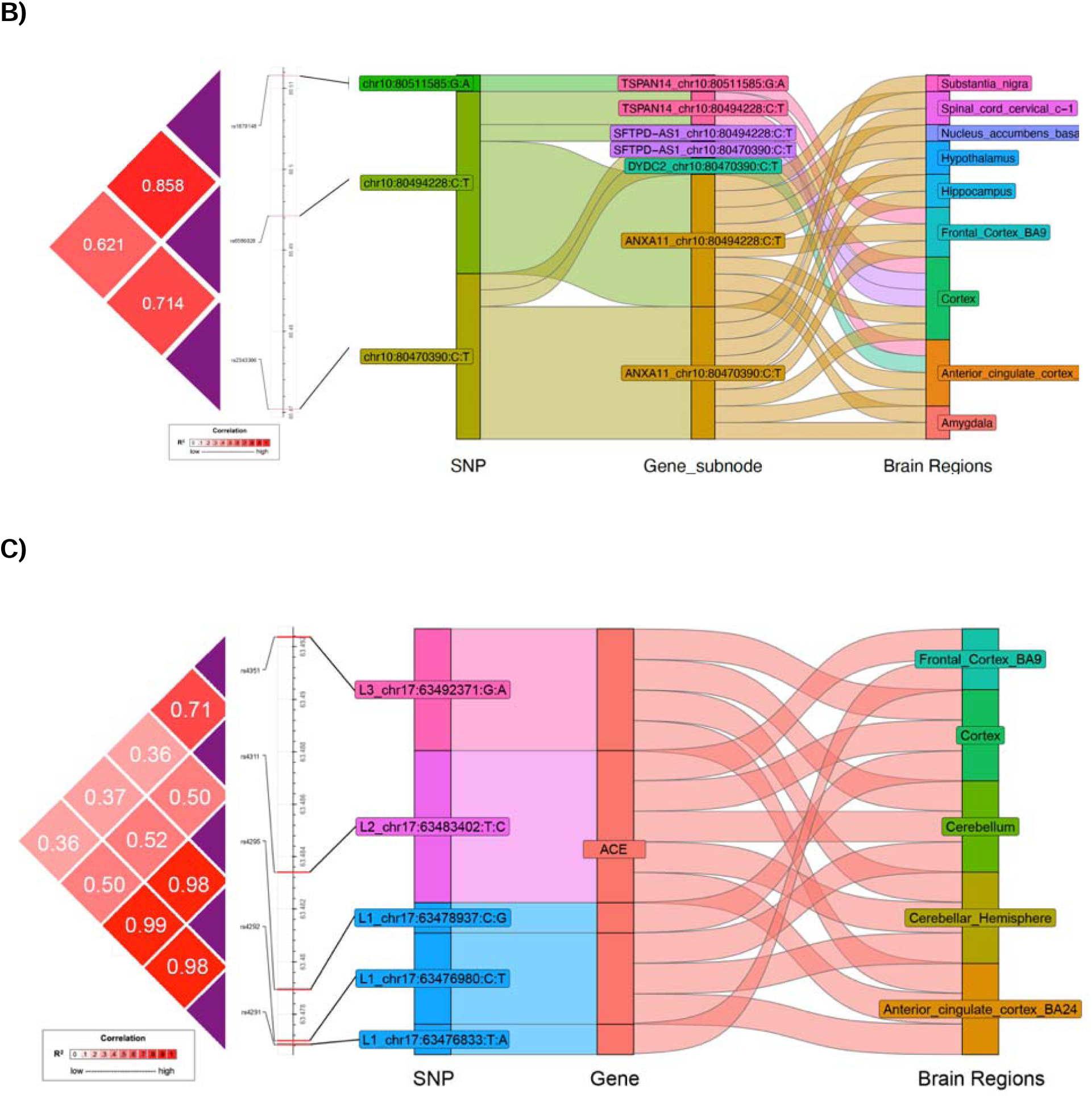
Colocalization results. **A)** A subset of significant colocalization results across 59 loci and 13 GTEx brain regions. **B)** All significant brain colocalization results of *TSPAN14*. **C)** Significant brain colocalization results from multiple independent loci inside the ACE region.

A single line of functional evidence is often considered insufficient to robustly implicate regulatory activity at a given locus. To address this limitation, we developed an unbiased confluent context identification strategy (*Methods: “Steps for unbiased confluent context identification”*) to integrate multiple lines of evidence to prioritize putative causal variants, genes, and V2G pairs for subsequent experimental validation. Of the 393 colocalized candidate variants, 68 (17%) overlap predicted HOMER^27^ TFBS (*Method: Transcription factor binding site (TFBS) disruption*). Among these, 15 (22%) and 23 (34%) fall within brain enhancers defined by ROADMAP^30^ and EpiMAP^31^, respectively (*Method: Enhancers overlap*). Overall, we identified 24 putatively causal variants (14 beyond the *APOE* locus) overlapping a brain eQTL, a brain enhancer (ROADMAP or EpiMAP), and a TFBS, forming 279 candidate tissue-V2G pairs involving 32 potential effector genes. Among non-*APOE* variants, 36% colocalized with >50% of GTEx brain tissues, with each variant interacting with an average of three (maximum 11) candidate effector genes. Notably, seven non-*APOE* variants did not affect the annotated (typically closest) genes in the original GWAS (**Table 1**)^5^. All selected colocalized candidate variants were common (non-reference allele frequency >0.05 in GWAS and 1000G). **Table 1** summarizes the 14 candidate regulatory variants (beyond *APOE*) identified by this unbiased confluent context identification approach. All eQTL effector genes and coloc results are detailed in **Supplementary Table 1**.

**Table 1:**
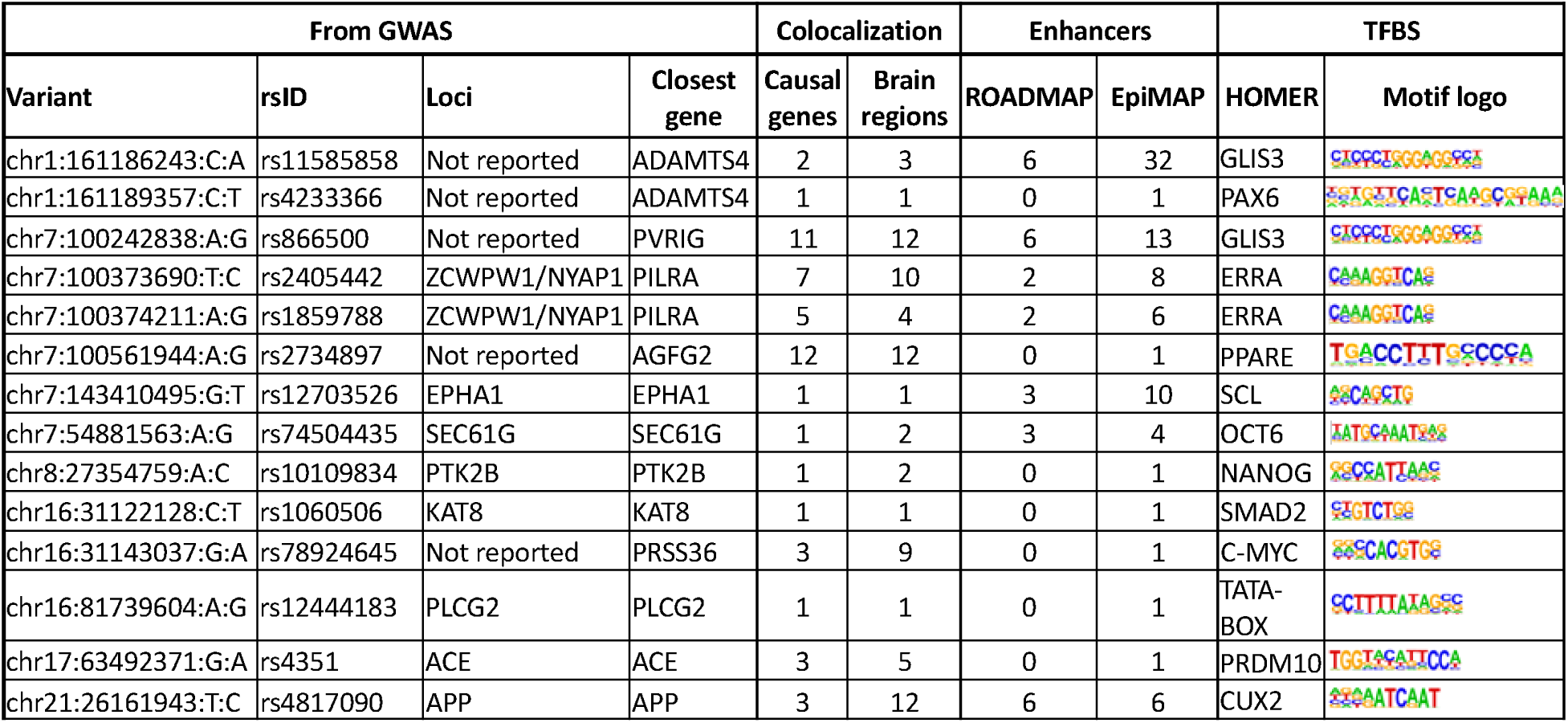
Genome-wide analysis of AD GWAS data implicates 14 candidate regulatory variants (outside chr19/*APOE*) with potential roles in brain tissue using the confluent context identification approach.

| From GWAS |  |  |  | Colocalization |  | Enhancers |  | TFBS |  |
| --- | --- | --- | --- | --- | --- | --- | --- | --- | --- |
| Variant | rsID | Loci | Closest gene | Causal genes | Brain regions | ROADMAP | EpiMAP | HOMER | Motif logo |
| chr1:161186243:C:A | rs11585858 | Not reported | ADAMTS4 | 2 | 3 | 6 | 32 | GLIS3 |  |
| chr1:161189357:C:T | rs4233366 | Not reported | ADAMTS4 | 1 | 1 | 0 | 1 | PAX6 |  |
| chr7:100242838:A:G | rs866500 | Not reported | PVRIG | 11 | 12 | 6 | 13 | GLIS3 |  |
| chr7:100373690:T:C | rs2405442 | ZCWPW1/NYAP1 | PILRA | 7 | 10 | 2 | 8 | ERRA |  |
| chr7:100374211:A:G | rs1859788 | ZCWPW1/NYAP1 | PILRA | 5 | 4 | 2 | 6 | ERRA |  |
| chr7:100561944:A:G | rs2734897 | Not reported | AGFG2 | 12 | 12 | 0 | 1 | PPARE |  |
| chr7:143410495:G:T | rs12703526 | EPHA1 | EPHA1 | 1 | 1 | 3 | 10 | SCL |  |
| chr7:54881563:A:G | rs74504435 | SEC61G | SEC61G | 1 | 2 | 3 | 4 | OCT6 |  |
| chr8:27354759:A:C | rs10109834 | PTK2B | PTK2B | 1 | 2 | 0 | 1 | NANOG |  |
| chr16:31122128:C:T | rs1060506 | KAT8 | KAT8 | 1 | 1 | 0 | 1 | SMAD2 |  |
| chr16:31143037:G:A | rs78924645 | Not reported | PRSS36 | 3 | 9 | 0 | 1 | C-MYC |  |
| chr16:81739604:A:G | rs12444183 | PLCG2 | PLCG2 | 1 | 1 | 0 | 1 | TATA-BOX |  |
| chr17:63492371:G:A | rs4351 | ACE | ACE | 3 | 5 | 0 | 1 | PRDM10 |  |
| chr21:26161943:T:C | rs4817090 | APP | APP | 3 | 12 | 6 | 6 | CUX2 |  |

To further explore the genetic architecture of gene regulation in the brain, we examined loci with multiple candidate causal variants and their effects on gene expression across brain regions. **Fig. 2B** highlights the significant colocalization results for the *TSPAN14* locus, where three candidate causal variants were associated with the expression of four genes across nine GTEx brain regions. **Fig. 2C** shows three independent signals L1, L2, and L3 (pairwise r^2^ = 0.501, 0.364 and 0.709, respectively) at the *ACE* locus each correlating with *ACE* gene expression across five GTEx brain regions. Together, these figures illustrate the complexity of genetic regulation, demonstrating how multiple independent variants can influence gene expression across diverse brain regions.

### Potential biological roles of the identified transcription factor binding motifs

We found that the candidate regulatory variants are predicted to disrupt binding sites for 12 transcription factors (TFs) based on PWM analyses (**Table 1**). Several of these TFs are involved in mechanisms related to AD. Notably, GLIS3 (GLIS family zinc finger 3) is the only motif strongly linked to tau and amyloid pathology through transcriptional regulation^32–34^. SMAD2 (Mothers against decapentaplegic homolog 2), a key intracellular protein in the SMAD family, transduces signals from TGF-β ligands and mediates cellular responses. TGF-β/SMAD2 pathway plays a complex role in AD, potentially affecting cell growth, differentiation, and immune responses^35^. Dysfunction in TGF-β signaling may lead to blood-brain barrier breakdown, and blocking TGF-β-SMAD2/3 signaling in peripheral macrophages has been proposed as a therapeutic strategy for AD^36^.

### In silico validation: identification of putative causal signals with further support

Outside the APOE region, we identified 42 candidate V2G pairs comprising 14 variants and 26 protein-coding genes that are likely functional in brain tissues or cell types. For additional in silico support, we leveraged independent FG datasets. Recent FG data from large consortia^37^, harmonized studies^38–41^, and individual publications^42,43^ provided complementary or orthogonal evidence for *in silico* validation. We processed these datasets using hipFG^44^ and integrated them into FILER^26^, including two QTL (MetaBrain^39^, eQTL catalogue^45^) and three EPI datasets (3DGenome^41^, 4DGenome^40^, Nott *et al.*^43^), standardizing all with metadata for efficient querying.

We first assessed the consistency of V2G pairs identified in brain regions across different data sources. Consistency in directionality means that a genetic variant’s effect on an eQTL (i.e., increasing or decreasing target gene expression) is the same across brain regions and data sources. Such consistency suggests shared genetic regulatory mechanisms across brain regions. However, eQTL directionality, represented by Z-scores, can vary between datasets due to (a) sampling differences, (b) QTL generation methods, or (c) statistical approaches. Since QTL datasets lack standardized presentation, harmonizing them with tools like hipFG can reduce these biases and improve interpretation.

In **Fig. 3**, the left panel shows hipFG-normalized Z-scores for each brain region (“System” in legend: Frontal Cortex, Limbic System, Basal Ganglia, Brain Stem) from two data sources (MetaBrain or eQTL Catalogue), along with average Z-scores across 13 GTEx brain regions (middle panel). The original, non-normalized Z-scores (**Supplementary Fig. 3**) show that 35 out of 42 V2G pairs had inconsistent directionality over >50% of the grouped brain regions. Strikingly, after Z-score normalization, inconsistencies dropped to only 6, an 83% improvement (McNemar’s Test, p<0.0001), as shown in **Fig. 3** (left panel).

**Fig. 3.**
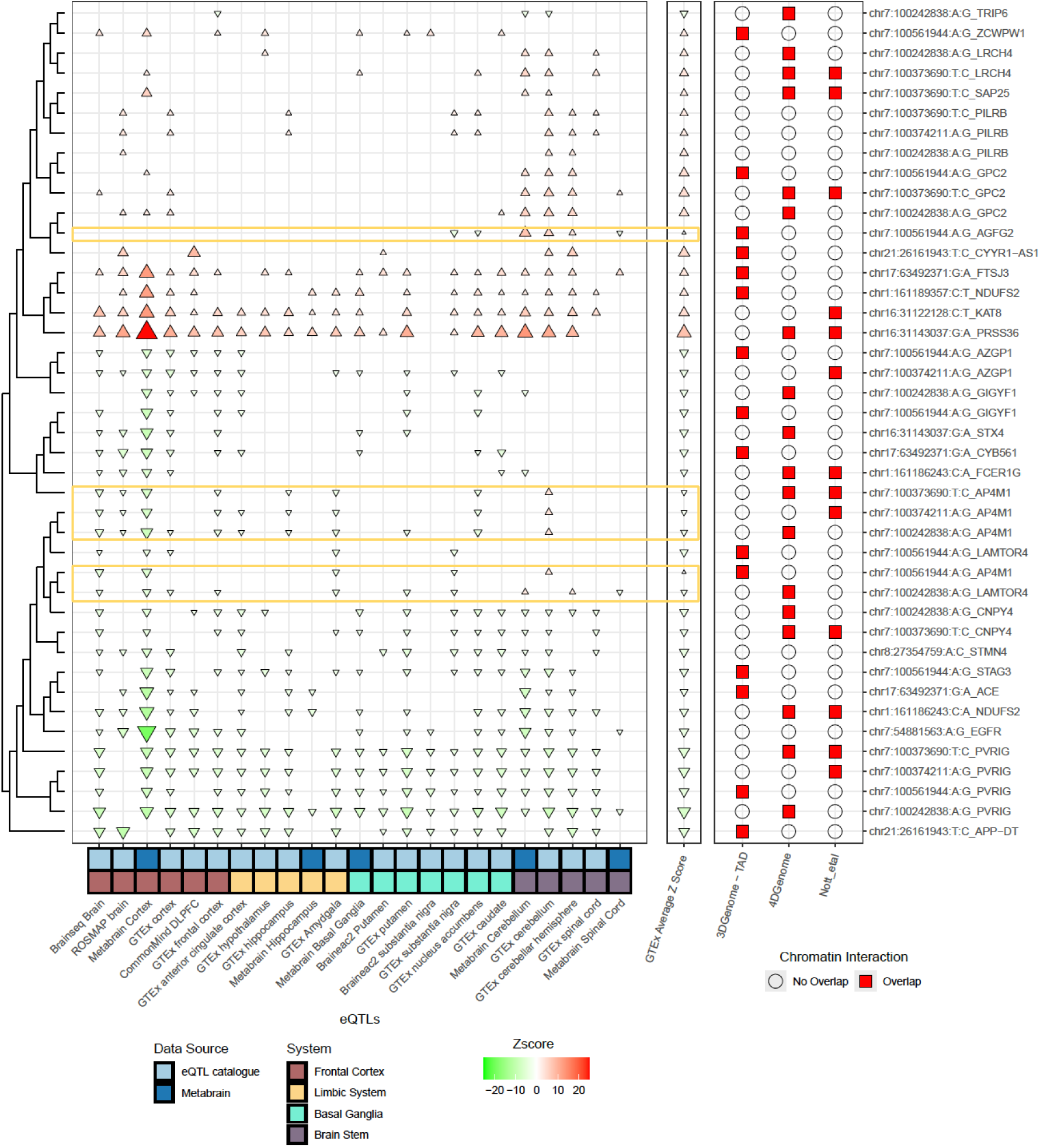
Comparison of the directionality of selected putative V2G pairs across QTL and EPI datasets used for *in silico* validation. All FG data were processed using hipFG, with effect directionality normalized. For both the left and middle panel, triangles pointing upwards (and in red) mean a positive Z-score, indicating that the alternative allele of a variant increases the gene expression, while those pointing downwards (and in green) mean the opposite effect. The size of the triangles represents the absolute value of the Z-score. The left panel shows the directionality (Z-scores) based on 23 QTL datasets from two data sources. The middle panel presents the average GTEx Z-scores, while the right panel displays the orthogonal support of EPI data for the V2G pairs (with a red square indicating the present of EPI for that particular data source). The six loci with inconsistent directionalities were shown in yellow boxes.

### A ranking system integrating in silico evidence on genes, variants and V2G pairs for selected putative causal signals

When an eQTL and an EPI co-occur for a given variant-to-gene (V2G) pair, it provides stronger evidence that the variant causally regulates gene expression^46,47^. To prioritize V2G pairs, we developed a ranking system based on four components: eQTL evidence (V2G_eQTL tier), EPI evidence (V2G_EPI tier), variant properties (V_tier), and gene expression (G_tier). Each tier was generated by integrating data from multiple sources (*Methods: Ranking of variant-gene pairs (Tier system)*). In more detail:

1) The V2G_eQTL tier assesses consistency of V2G pairs across gene expression datasets (beyond just the GTEx): a) directionality across non-GTEx eQTLs (MetaBrain^39^, eQTL Catalogue^45^), b) presence of non-GTEx brain eQTLs, c) consistency between non-GTEx and GTEx brain region eQTLs, and d) consistency across brain regions.
2) The V2G_EPI tier checks for V2G pairs in brain EPIs profiled in brain from 3DGenome^41^, 4DGenome^40^, or Nott *et al.*^43^.
3) The V_tier evaluates variant characteristics: linked to active gene regulation marks (active histone marks), located in open chromatin regions, deleterious to regulatory region function (CADD scores)^48^, predicted functional by ENCODE data (RegulomeDB2 ranking)^49^, and genome-wide significant in GWAS (GWAS Catalog^50^, NIAGADS GenomicsDB^51^).
4) The G_tier considers gene relevance and expression: previously linked to LOAD (AMP-AD Agora^37^ gene nomination), expressed in brain regions or cell types (HPA)^52,53^.

This structured approach helps prioritize V2G pairs based on robust, multi-source evidence.

The V2G_eQTL and V2G_EPI information are presented in **Fig. 3** while overall tier rankings are displayed in **Fig. 4**. The top-ranked V2G pair was chr1:161186243:C:A_*NDUFS2* (tier 11), followed by four pairs at tier 9 (chr7:54881563:A:G_*EGFR*, chr1:161186243:C:A_*FCER1G*, chr7:100373690_*PVRIG,* and chr17:63492371:G:A_*ACE*). These five pairs are hypothesized to have the greatest likelihood of functional study success.

**Fig. 4.**
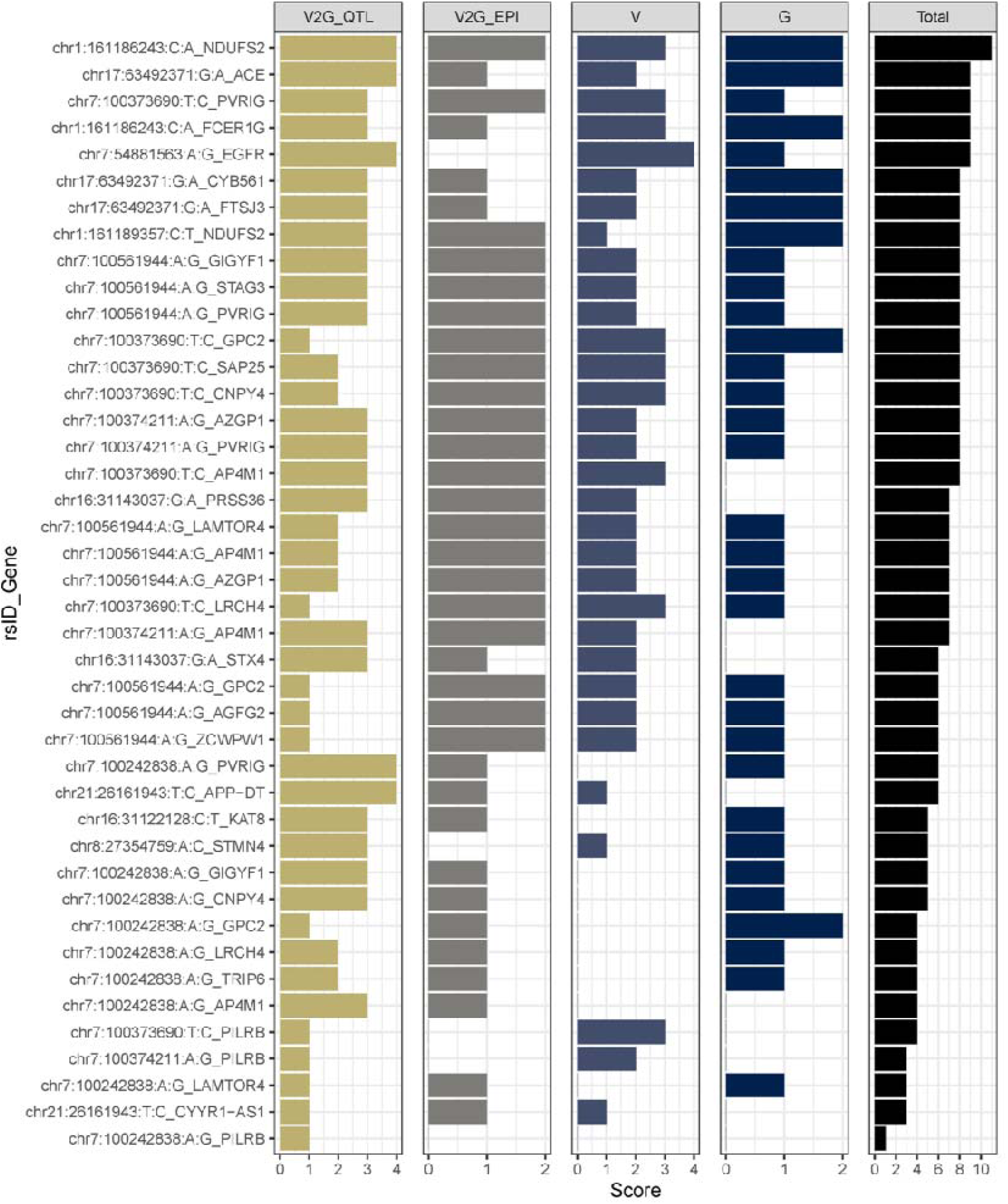
Rankings (V2G_eQTL tier, V2G_EPI tier, V_tier, G_tier and overall tier) of the 42 V2G pairs identified in this study.

### Functional validation: an enhancer region harboring rs74504435 influences EGFR expression

We selected the second-highest-ranking V2G pair - rs74504435-*EGFR -* for further validation due to two main reasons. The LOAD risk allele at rs74504435 is associated with increased *EGFR* expression in multiple eQTL datasets (**Fig. 5a**), suggesting *EGFR* as a promising candidate for therapeutic targeting with existing *EGFR* inhibitors, This pair ranked highest in both the V2G_eQTL and V tiers, indicating robust support from diverse eQTL datasets and consistent functional annotations predicting a regulatory (enhancer) function for rs74504435 (**Fig. 5a**). **Fig. 5a** visualizes this V2G pair using FILER tracks and Bellenguez GWAS summary statistics. rs74504435 regulates *EGFR* in 4 eQTLs: ROSMAP DLPFC, CommonMind DLPFC, GTEx Frontal Cortex and GTEx cortex. It is also located within four brain-related enhancers from chromHMM and EpiMAP.

**Fig. 5a.**
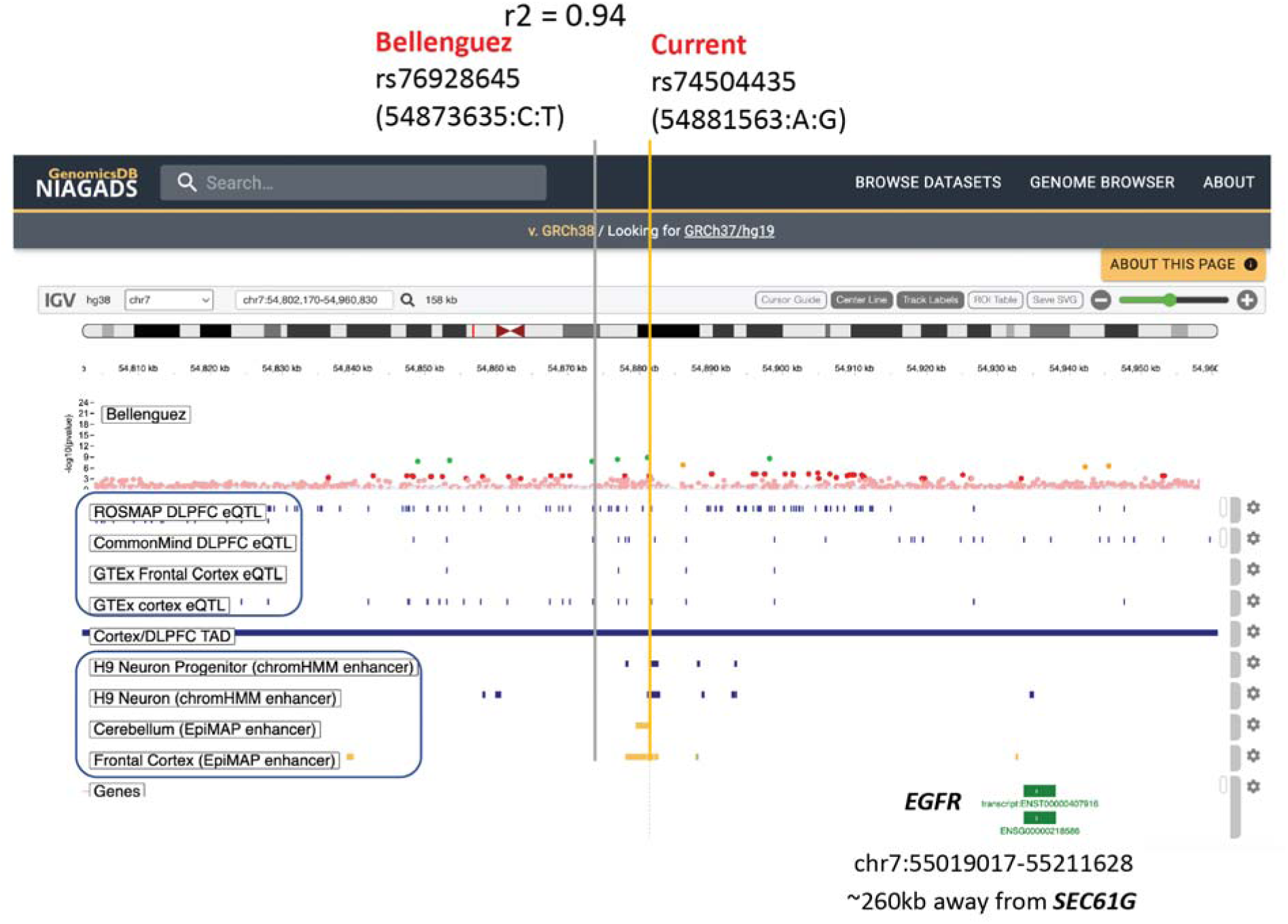
Genome browser plot showing the functional annotation of chr7:54881563:A:G (rs74504435) and *EGFR* (GRCh38) using FILER tracks at the discovery phase, including enhancers from chromHMM and EpiMAP, eQTLs from GTEx and eQTL catalogue. Bellenguez GWAS identified the sentinel SNP rs76928645 (chr7:54873635:C:T) (*p*=1.6×10^-10^), in high LD (r^2^=0.94) with our variant of interest rs74504435, which is annotated to reside in a brain enhancer in four different sources. Plot is generated using NIAGADS genomicsDB.

To investigate a potential regulatory role for rs74504435 on *EGFR* expression, we next validated this V2G pair by leveraging our collection of promoter-focused Capture C, ATAC-seq, and RNA-seq datasets from human brain-relevant cell types^54–56^, as well as Hi-C data from iPSC-derived astrocytes, microglia, neurons and oligodendrocytes^57^. Via the ATAC-seq dataset, We observed that rs74504435 lies within open chromatin in several brain-relevant cell types, including iPSC-derived cortical neural progenitors and neurons^55^, primary astrocytes^56^, and iPSC-derived microglia^54^. We also observed a chromatin conformation capture contact (Capture C data) between this variant and *EGFR* in iPSC-derived cortical progenitors and neurons, primary astrocytes, and the microglial cell line HMC3. Using our RNA-seq datasets, we observed that *EGFR* was indeed expressed in these cell types^54–56^. From the Hi-C data, we observed that interactions between rs74504435 and *EGFR* exist in iPSC-derived neurons and oligodendrocytes. These findings are illustrated in **Fig. 5b**.

**Fig. 5b.**
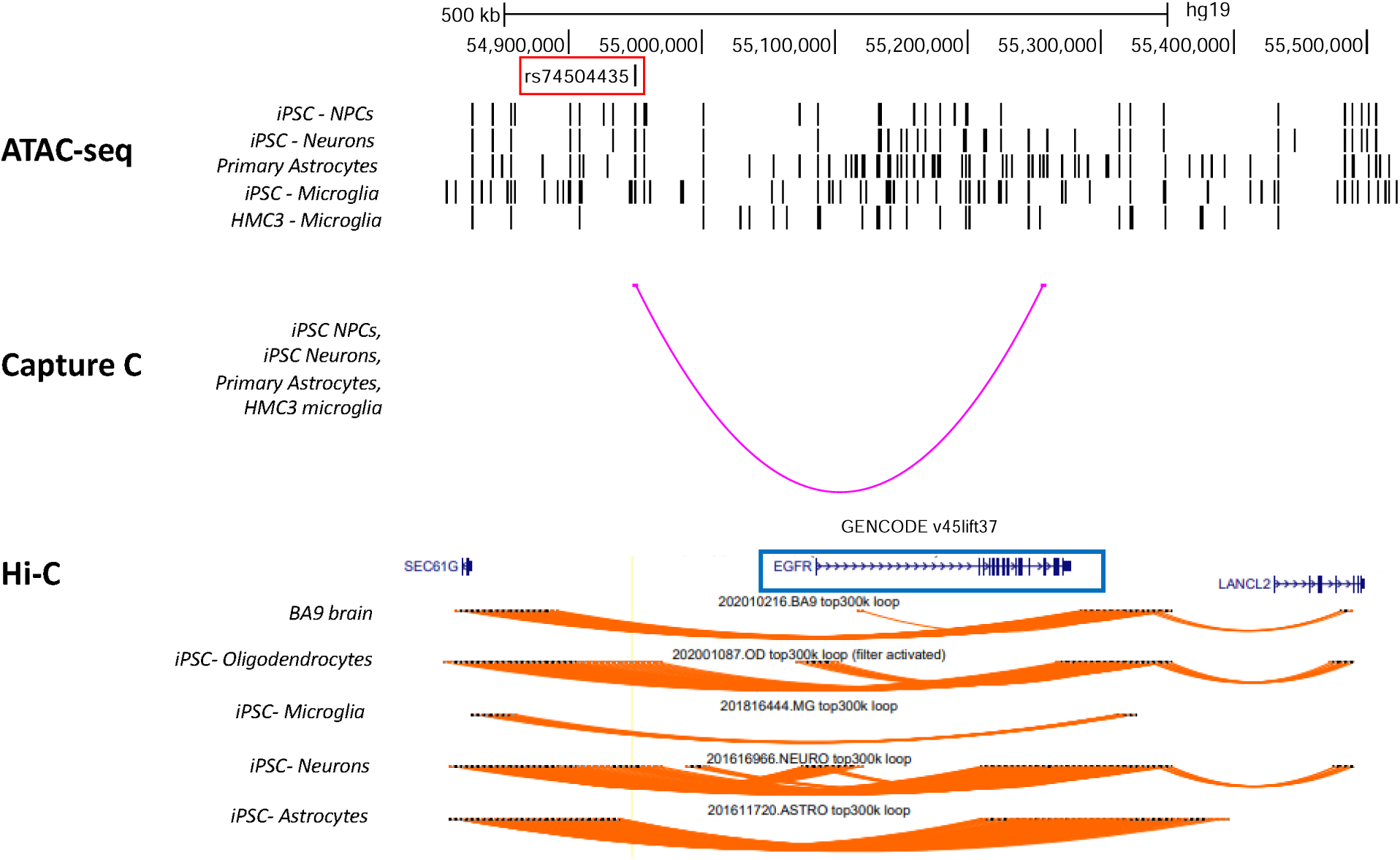
ATAC-seq, promoter-focused Capture-C, and Hi-C data in brain-relevant cell types showing chromatin state and looping between chr7:54881563:A>G (rs74504435) and *EGFR*. rs74504435 (highlighted by a yellow line) resides in open chromatin in iPSC-derived neural progenitors, neurons, and microglia, and in primary astrocytes. It is involved in a chromatin loop with *EGFR* in iPSC-derived neurons, primary astrocytes, and the microglial cell line HMC3 (promoter-focused Capture C data); and in iPSC-derived neurons and oligodendrocytes (Hi-C data).

To experimentally validate the regulatory role for the region harboring rs74504435 and its influence on *EGFR* expression, we leveraged CRISPR interference (CRISPRi). We engineered the human microglial cell line HMC3 to stably express dCas9-KRAB (tagged with GFP). We transduced this line with lentivirus containing one of three sgRNAs targeting this region (E1, E2, and E4) or two control non-targeting sgRNAs (tagged with mCherry). After double selection for the presence of the guides and the dCas9-KRAB by FACS, we performed qPCR to assess *EGFR* levels. We found that two out of three targeting guides led to a consistent and significant decrease in *EGFR* expression levels compared to controls (one-way ANOVA *p*=0.0002). E1 significantly decreased *EGFR* levels by 47% compared to the mean of the non-targeting controls (Tukey test *p*=0.0004), and E2 by 27% (*p*=0.04). E4 did not affect *EGFR* levels. Non-targeting guides did not show any effect when compared to a no guide control. These results are shown in **Fig. 5c**.

**Fig. 5c.**
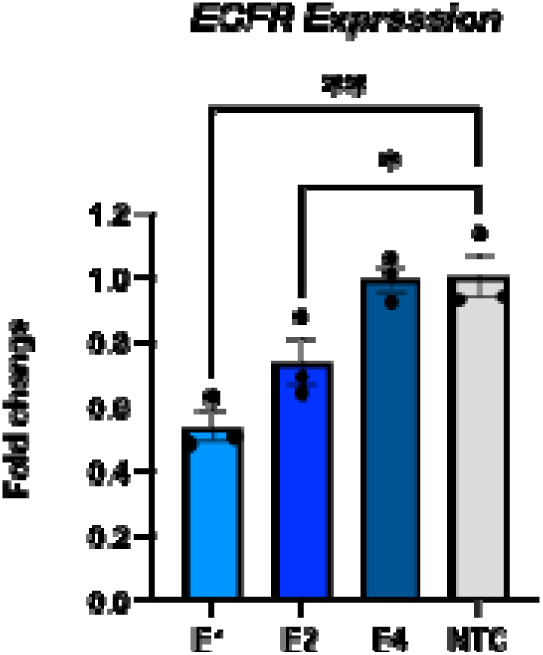
CRISPRi results in human microglia cell line. We performed CRISPRi in a human microglial cell line (HMC3) expressing dCas9-ZIM3-KRAB using lentiviral delivery of three sgRNA guides targeting the rs74504435 region (E1, E2, and E4) and two non-targeting guides (NTC: mean of control guides). Bar plots show the mean *EGFR* relative expression compared to a no guide control as assessed by qPCR; error bars are SEM; N=3. Statistical analysis via one-way ANOVA followed by Tukey test, ** *p*<0.001; * *p*<0.05.

Overall, our results support the hypothesis that the AD-associated variant rs74504435 is located within an enhancer region that regulates *EGFR* expression levels.

## DISCUSSION

Functionally characterizing non-coding AD GWAS loci is crucial for successful drug target discovery, and FG datasets can aid in this challenging task. However, FG data are often sparse and unharmonized, which hinders progress in this field. Here, we leveraged hipFG, a tool that integrates FG data into FILER, and selected brain-profiled FG data to systematically validate non-coding AD signals for their regulatory potential. Our new framework combines confluent FG evidence and directionality checks to prioritize V2G pairs for functional validation.

Starting with AD GWAS variants from Bellenguez *et al.*, 2022^5^, we defined independent loci using 1000G^28^ LD structure. For each locus, we evaluated functional contexts, conducted Bayesian co-localization of GWAS and eQTL signals, and identified putative causal variants, genes, and V2G pairs. We performed *in silico* validation using independent assays and data sources, followed by experimental validation with promoter-focused Capture-C, ATAC-seq, and CRISPRi. This approach identified five plausible V2G pairs with highest tiers: chr1:161186243:C:A_*NDUFS2* (tier 11), chr7:54881563:A:G_*EGFR* (tier 9), chr1:161186243:C:A_*FCER1G* (tier 9), chr7:100373690_*PVRIG* (tier 9), and chr17:63492371:G:A_*ACE* (tier 9). We successfully validated one of these pairs and its regulatory effect in a human microglia cell line.

Unlike prior post-GWAS methods, which rely on pre-selected top GWAS variants^11,25^, our framework utilized full GWAS summary statistics. We implemented QC steps to normalize genetic data against FG datasets in FILER, enabling a more systematic and comparable analysis of potential causal variants, genes and V2G pairs. Our tiering system, which integrates support from eQTL, enhancer-promoter interactions, variant effects, and gene/protein expression, enabled improved prioritization of V2G pairs.

By leveraging full GWAS summary statistics and LD expansion in post-GWAS analyses, we uncovered additional candidate causal signals that were missed in previous studies^5^. When we compared the number of colocalization signals (the first step in our pipeline, GTEx brain data alone) to analyses that did not perform LD pruning or expansion^11^, we found that using the full summary statistics, rather than only top variants, yielded 4.5 times more candidate variants (9,144 vs. 2,026), 5 times more unique colocalized variants (2,040 vs. 408), and twice as many candidate target genes (1,529 vs. 762) across all GTEx data.

Using eQTL data for colocalization explained only a small fraction of the GWAS signals^58^. In our approach, we did not restrict analysis to signals with EPI support. Instead, we first prioritized eQTL colocalization and then sought additional *in silico* validation. 42 eQTLs were validated using independent eQTL datasets, and 37 out of 42 also had EPI support. EPI data can help prioritize V2G pairs when eQTL signals are weaker (e.g., loci chr7:100374211:A:G_CNPY4 and chr7:100373690:T:C_AP4M1). However, V2G pairs supported by both types of evidence are not always superior, as discrepancies may arise from biological differences or the uneven availability of FG assays, which limits cross-cell-type comparisons. More cell-type-specific assays, currently unavailable, could further improve V2G pair identification.

Using CRISPRi in a microglial cellular setting, we successfully validated *EGFR* as a target gene whose regulation is influenced by the AD variant rs74504435. The *EGFR* (Epidermal Growth Factor Receptor) gene product is a receptor tyrosine kinase that controls cell proliferation, survival, differentiation, and inflammation. In AD, it has been connected to disease progression^59^, with elevated levels associated with increased Aβ plaque formation. Additionally, *EGFR* inhibition modulates neuroinflammation and cognitive function in AD animal models^60^.

Interestingly, we found evidence supporting a role for the rs74504435-EGFR V2G pair in multiple brain-relevant cell types: 1) rs74504435 resides in open chromatin regions of neurons, microglia, and astrocytes in ATAC-seq data; 2) rs74504435 contacts the 3’UTR region of *EGFR* in promoter-focused Capture-C data generated from microglia, neurons and astrocytes; 3) rs74504435 also contacts the *EGFR* promoter in Hi-C data derived from iPSC-derived oligodendrocytes and neurons; 4) rs74504435 is associated with *EGFR* expression in single-nucleus RNA-seq data from astrocytes (*p*=3.2×10^-24^, Z-score=-12.53) and oligodendrocyte progenitor cells (*p*=0.003, Z-score=-2.95) derived from the dorsolateral prefrontal cortex in ROSMAP samples (Supplementary Fig 4)^61^. As a consequence, we conclude that *EGFR*’s functional role in AD likely involves several cell types, and further research is required to investigate whether the underlying mechanisms are distinct or shared across different brain cell types and whether pathogenesis is driven by a specific cell type.

In conclusion, by combining an unbiased, confluent context identification framework, *in silico* V2G pair validation using QTL directionality, and a comprehensive scoring system that considers variant, gene, and V2G pair effects, we identified 42 AD-associated V2G pairs. The FILER-curated brain FG datasets and hipFG-harmonized FG data were instrumental to this success. Among the top findings, five V2G pairs achieved tier 9 or higher, and we demonstrated that AD-associated variant rs74504435 (chr7:54881563:A:G) resides in a regulatory region influencing *EGFR* expression, as validated by promoter-focused Capture C and CRISPRi functional experiments. Given that *EGFR* inhibitors are already approved for cancer therapy, *EGFR* could represent a promising candidate for repurposing as a therapeutic target for LOAD. Our framework and results provide valuable insights for future AD research.

## METHODS

### Description of the AD GWAS

We analyzed Stage 1 genome-wide summary stats from Bellenguez *et al.*, 2022^5^, (downloaded from GWAS catalog http://ftp.ebi.ac.uk/pub/databases/gwas/summary_statistics/GCST90027001-GCST90028000/GCST90027158/). This dataset aggregates samples from International Genomics of Alzheimer’s Project (IGAP), with the inclusion of cohorts from European Alzheimer’s & Dementia BioBank (EADB), Alzheimer’s Disease Genetics Consortium (ADGC), and others, including clinically defined AD cases/controls and UK Biobank dementia samples, totaling 788,989 individuals with an effective sample size of 382,472. It contains 19,767,628 SNPs and 1,333,486 indels; our analysis focused on SNPs. For details, refer to the original publication^5^.

### Identifying regions of interest for downstream analyses

We performed LD pruning using the 1000 Genomes Phase 3 EUR reference panel on all genome-wide significant variants (*p*<5×10^-8^) identifying pairwise-independent tag variants (r^2^<0.7) using a 500kb window. For each tag variant, we defined an analysis region by including variants that 1) are in LD with the tag variant (r^2^>=0.7), 2) are within 1M basepairs (bps), and 3) are within 1,000 variants of the tag. These variants, along with the tag variants, are considered candidate regulatory variants. Each analysis region is bounded by the outermost variants in LD with the tag. We note that not only the candidate variants, but all variants within these regions, even if some have no association with AD, will be included in colocalization analyses.

### Characterization

#### Genome partition analysis

Variants were categorized into different genomic categories using the UCSC knownGene^62,63^, UCSC RepeatMasker^63,64^ and GENCODE v43 lncRNA annotations^65^ for the GRCh38/hg38 genome build. The 5′ UTR exons and introns, exons, introns, and 3′ UTR exons and introns were extracted from the knownGene annotation for each protein-coding gene. Promoter annotations were defined as 1000 bps genomic regions upstream of the transcription start site. To create a hierarchical genomic partition into disjoint 5’ UTR exon, 5’ UTR intron, 3’ UTR exon, 3’ UTR intron, promoter, exonic, intronic regions (in this order), each region set was obtained by subtraction of the merged regions higher in the genomic hierarchy. For example, starting from merged 5’ UTR exonic regions, distinct 5’ UTR intronic regions were obtained by subtraction of 5’ UTR exonic regions from the merged 5’ UTR intronic regions, and 3’ UTR exon regions were obtained by subtracting both 5’ UTR exonic and intronic regions.

During analysis, GWAS variants were then assigned to mutually exclusive genomic element annotations using the created hierarchy: 5′ UTR exon > 5′ UTR intron > 3′ UTR exon > 3′ UTR intron > promoter > mRNA exon > mRNA intron. Overlaps, if any, with repeat element annotations (e.g., SINE, LINE) were also reported for all variants. Variants overlapping any of GENCODE lncRNA annotations were additionally classified into lncRNA exonic and/or lncRNA intronic variants. A variant not overlapping with any class of elements above (mRNA, lncRNA, repeat) was classified as intergenic.

#### Functional Genomic Annotations

Genomic annotations from the FILER (functional genomic database which contains harmonized genomic annotation data across >30 primary data sources^26^) were used. 140 of these are used in the **“Characterization”**, i.e. discovery phase (including genome partition analyses, colocalization analyses, and unbiased confluent context identification) while 34 are used in “In silico validation”. See **Supplementary Table 2** for details. In the discovery phase, the datasets included 1) fundamental genome annotations and reference variant information (dbSNP^66^, GENCODE gene annotations^65^), 2) genome-wide HOMER^27^ transcription factor binding tracks. 3) 140 brain-related FILER tracks (tissues and cells only) for variant characterization, including enhancers (10 from ROADMAP^30^, 55 from EpiMAP^31^), QTLs (26 from GTEx v8^29^), and epigenetics (49 from ENCODE^67^).

#### Variant-Gene (V2G) pair identification (colocalization)

We hypothesized that putative causal variants affect gene expression in a cis-regulatory manner. By aligning GWAS and eQTL signals via colocalization, we can identify genes most likely affected by disease-associated variants. Using the COLOC R package v5.2.3^68^, we performed Bayesian colocalization on 9,058 candidate variants against nominally significant eQTLs from 13 brain tissues in the GTEx v8 dataset. We defined a V2G pair as colocalized if: 1) the candidate variant is the most likely causal variant in the locus (coloc_SNP.H4.abf > 0.5), 2) the posterior probability for colocalization is > 0.7 (coloc_PP.H4.abf), and 3) the locus contains more than one variant.

#### Transcription factor binding site (TFBS) disruption

HOMER (Hypergeometric Optimization of Motif EnRichment)^27^ is a custom motif database derived from high-quality ChIP-Seq data. A positional weight matrix (PWM) represents transcription factor (TF) DNA binding specificities. The delta PWM score (difference between reference and alternate alleles) estimates binding activity changes due to nucleotide variation. A candidate causal variant is selected for next steps if it disrupts a TF binding site with a delta PWM score >|2| for any TF.

#### Enhancers overlap

Enhancers are DNA regulatory elements that activate gene transcription by forming chromatin loops to interact with target genes in a cell-type-specific manner. Databases like ROADMAP^30^ and EpiMAP^31^ catalog enhancers across various cell types and tissues. A candidate causal variant is considered potentially regulatory if it overlaps a brain enhancer found in either ROADMAP^30^ or EpiMAP^31^.

#### Steps for unbiased confluent context identification

We define a putative causal V2G pair as one with strong colocalization (***SNP-Gene pair identification (colocalization)***). The associated genes are considered putative causal genes. The putative causal variant is predicted to disrupt a TFBS (***Transcription factor binding site (TFBS) disruption***) and overlap a tissue- or cell-type-specific enhancer (***Enhancers overlap***) in any brain FG data. To identify which AD genetic signals may function in the brain-specific confluent context, we included all relevant tracks in FILER. We then required the colocalization tissue context to match the enhancer’s, forming the final set of putative causal V2G paris. A pair is excluded if it fails to meet any of these criteria.

### In silico validation

#### Harmonizing in silico datasets by hipFG

In the validation phase, selected brain tracks include chromatin interactions (5 from 3DGenome^41^, 1 from 4DGenome^40^, 2 from Nott *et al.* 2019^43^), QTLs (18 from eQTL Catalogue^45^, 4 from MetaBrain^39^, 4 from MiGA^42^). These tracks were generated from primary tissues/cell types but not cell lines. Each dataset was processed using hipFG (Harmonization and Integration Pipeline for Functional Genomics)^44^, an automated pipeline that standardizes, indexes, and integrates diverse functional genomics data (e.g., EPI, genomic intervals, QTLs) for scalable, searchable analysis.

#### In silico validation on candidate variant-gene pairs and genes

We validate selected V2G pairs and genes *in silico* using a set of independent FG resources (**Fig. 1, “In silico validation**”). Validation requires evidence from at least one, not all, of the following categories.

1) V2G pairs

a. TAD - we used the TADs shared by the 3DGenome^41^. A V2G pair is *in silico* validated if it overlaps with both anchors of the interaction.
b. EPI– We included the EPI data profiled by PLAC-seq, 3C, 4C-Seq, 5C, Hi-C, ChIA-PET and promoter-focused Capture-C from Nott *et al.*^43^, 3DGenome^41^, and 4DGenome^40^. Only brain related datasets were used. A V2G pair is *in silico* validated if it overlaps with both anchors of the interaction.
c. Bulk tissue eQTL – Brain tissue / region related eQTLs from the eQTL catalogue^45^ and the MetaBrain^39^ from four brain regions were included. A V2G pair is *in silico* validated if it i) overlaps with; ii) contains the same effect allele; and iii) carries the same effect direction on the same gene against any eQTL profiled in any of these resources.
2) Genes

a. Biological context – information from Human Protein Atlas (HPA)^52,53^. A gene is *in silico* validated if it is found in the HPA to be a protein-coding gene expressed in any of the brain regions or cell types.
b. Disease specificity – information from Agora AMP-AD platform^37^. A gene is *in silico* validated if it is within the nominated list of genes.

#### Independent regulatory evidence and potential functions of selected variants

In addition to functional evidence from *in silico* validation, annotations from other sources can further support a variant’s regulatory or functional potential. As with previous analyses, evidence from any (not all) of the following categories is sufficient for confirming a variant as *in silico* functional.

1) Active histone marks (H3K27ac) from ENCODE^67^ are used. A variant is considered functional if it significantly overlaps any active histone mark peak (q-value <5%).
2) Open chromatin regions were analyzed using ATAC-seq data from ENCODE^67^. A variant is considered functional if it falls within an ATAC-seq peak.
3) We used the Combined Annotation Dependent Depletion (CADD) score ^48^ and the RegulomeDB2 score ^49^ for variant effect prediction. CADD assesses variant deleteriousness, with higher scores indicating greater impact. RegulomeDB2 annotates variants by intersecting their positions with functional genomic assays and assigns a heuristic ranking for regulatory potential. A variant is considered functional if it has a CADD score >10 or a RegulomeDB2 score of 1a,b,c,d,e.
4) We used the GWAS Catalog ^50^ and NIAGADS GenomicsDB ^51^ to assess whether a putative causal variant is linked to AD-related traits, indicating functionality. The GWAS Catalog includes variant-trait associations from >130,000 GWASs across >18,000 traits (as of February 2025), while NIAGADS GenomicsDB is an interactive AD genetics database with 476.9K annotated variants from >80 AD GWASs. A variant is considered functional if in addition to Bellenguez *et al.* (2022) AD GWAS the variant is also associated with a phenotype in any other AD GWAS.

#### Ranking of variant-gene (V2G) pairs (Tier system)

Given the large number of variant-gene pairs identified after the confluence analyses, it remains challenging for wet-lab scientists to prioritize pairs for further functional work. We introduced a tiered system that integrates evidence from eQTLs (V2G_eQTL tier), EPIs (V2G_EPI tier), variants (V_tier), and genes (G_tier), each combining data from multiple sources (*Methods: Ranking of variant-gene pairs (Tier system)*).

1) V2G_eQTL tier assesses: a) directionality across non-GTEx eQTLs (Metabrain^39^, eQTL Catalogue^45^), b) the presence of non-GTEx brain eQTLs, c) consistency between non-GTEx and GTEx brain region eQTLs and d) a negative, consistent Z-score across brain regions. Each of these criteria will be assigned a score of “1”. The V2G_eQTL tier score (the sum of these four criteria) ranges from 1 to 4.
2) V2G_EPI tier checks if a V2G pair appears in brain EPIs from 3DGenome^41^, 4DGenome^40^, or Nott *et al*^43^. A score of 1 will be given to each presence. The V2G_EPI tier ranges from 0 to 2.
3) V_tier evaluates variant properties, including overlap with active histone marks, open chromatin, CADD scores^48^, RegulomeDB2 ranking^49^, and statistical significance in GWAS (GWAS Catalog^50^, NIAGADS GenomicsDB^51^). A ‘functional’ definition in each of this category will be given a score of 1. The V_tier ranges from 0 to 4.
4) G_tier considers gene nomination by AMP-AD Agora^37^ and expression in brain regions or cell types (HPA)^52,53^. A score of 1 will be given to each presence. The G_tier ranges from 0 to 2.

The overall tier for V2G pair is the sum of all four components. The overall tier ranges from 1 to 11.

### Functional validation

#### Hi-C data generation from iPSC cells

To understand the potential function of the intergenic SNP rs74504435, we examined chromatin interactions using Hi-C analysis in iPSC-derived astrocytes, microglia cells, oligodendrocytes and neurons^57^. *In situ* Hi-C library was prepared using the protocol adapted from Rao *et al.*^69^. For each library, 450∼550 millions of paired-end reads at 150bps length were obtained. Sequencing data were processed using BWA^70^ to map each read end separately to GRCh19 reference genomes. Duplicate and non-uniquely mapped reads were removed. For each library, over 270 million of non-redundant, uniquely mapped, paired reads were used for further analysis. For robust enhancer-promoter interaction mapping, Chromatin loops were called using HiCorr^71^ to correct bias and LoopEnhance^72^ to remove noise.

#### Chromatin interaction analysis

We queried our existing genomic datasets including high-resolution promoter-focused Capture C, ATAC-seq and RNA-seq from brain relevant cell types (iPSC-derived neural progenitors and neurons^54^, iPSC-derived microglia and the human microglia cell line HMC3^55^, and primary astrocytes^56^) to assess whether candidate variants were residing in open chromatin regions and contacting the promoter of an expressed gene.

#### EGFR sgRNA design

The genomic coordinate location for rs74504435 was obtained using the UCSC genome browser (build GRCh19). This genomic coordinate for rs74504435 plus and minus 200bps was then entered into the software CHOPCHOP^73^ to generate a table of possible single guide RNAs (sgRNAs) for CRISPRi (repression) using Cas9. The Cas-OffFinder software^74^ was used to access the off-target mismatches of the possible sgRNAs generated from CHOPCHOP. After assessing the efficiency and off-target mismatches, three sgRNAs targeting rs7450443 (E1, E2, E4) were selected. Two non-targeting control guides (N2 and NTC3, Millipore Sigma) were used as negative controls for the CRISPRi experiments. As a positive control, we utilized a guide targeting an enhancer of the gene *TSPAN14*, since we previously validated this construct for CRISPRi experiments in HMC3.

#### Cloning the sgRNAs in a lentiviral plasmid

We leveraged a lentiviral vector created in the Chesi lab (SL33 Lenti-sgRNA(Tp2)- mCherry) to generate the backbone and the insert required for NEB HiFi DNA Assembly cloning. This lentiviral vector contains a U6 promoter driving the expression of one sgRNA; it also contains an sgRNA scaffold region and mCherry as a selection marker. To clone the sgRNA of interest into this vector, forward and reverse primers were designed and ordered through Azenta. The forward primer included: the sequence of the sgRNA of interest, complementary bases to the sgRNA scaffold, and overhanging bases (complementary to the vector - which is needed for Hifi cloning). The reverse primer was designed in such a way that when used with the forward primer in PCR, the amplicon would be the HiFi insert, i.e. the sequence of the sgRNA of interest, the sgRNA scaffold, and overhanging bases complementary to parts of the HiFi backbone. The SL33 Lenti-sgRNA(Tp2)-mCherry vector was digested with restriction enzymes XhoI and BsrG1 and ran on a gel to isolate and extract the HiFi backbone. The forward and reverse primers were used in a PCR cycle with the SL33 Lenti-sgRNA(Tp2)- mCherry as template in order to obtain the HiFi insert. The backbone and the insert were combined in HiFi cloning to generate the respective sgRNA lentiviral plasmids. The plasmids were then transformed using NEB 5 alpha competent bacteria. After colony picking and miniprep, plasmids were submitted to Plasmidsaurus for Nanopore sequencing for validation. We cloned three sgRNAs designed to target rs74504435 (E1, E2, E4) and two non-targeting sgRNAs (N2 and NTC3).

#### Virus generation

On day 0, 400,000 HEK 293T cells per well were plated on PDL coated 6-well plates. On day 1, cells were transfected with the respective sgRNA lentiviral plasmid in addition to envelope (Addgene plasmid #12259) and packaging (Addgene plasmid #12260) plasmids using the Lipofectamine 3000 reagent. On day 2, complete media changes were performed. On day 4, media from the HEK Cells (Virus Day Two) were collected, filtered through a 0.45-micron filter, and aliquoted and frozen/stored in the −80C.

#### Transducing HMC3 helper line cells

We generated a CRISPRi helper line by transducing the human microglial line HMC3 (ATCC #CRL-3304) with a lentiviral vector encoding Zim3 Krab dCas9 and GFP as a selection marker (Addgene plasmid #188778). This line (HMC3-zim3) was plated in a 6 well format - 200,000 cells per well on day 0. On day 1, cells were transduced with Day Two virus, and polybrene was used to aid in transduction efficiency. Due to the toxic nature of polybrene if cells are exposed to it for too long, a complete media change was performed 18 hours later (day 2). On the same day, cells were later moved from the 6 well format to a 100 mm plate in order to expand them for fluorescence activated cell sorting (FACS). After cells reached about 80-100% confluency on the 100 mm plate, FACS was performed on these transduced cells as well as naïve HMC3 cells (used as a baseline for fluorescence) at the Flow Cytometry Core Laboratory at CHOP. Double positive cells were selected, i.e. the top 50% of cells that had both GFP fluorescence (indicating that these cells have dCas9) and mCherry fluorescence (indicating that these cells had the sgRNA lentiviral vector) were chosen. After recovering from flow sorting, cells were cultured and expanded until there were enough to perform qPCR experiments.

#### qPCR

qPCR primers for *EGFR* (spanning exons 7-9), *TSPAN14* (spanning exons 8-9), and *GAPDH* (spanning exons 2-3) were ordered through IDT. After cells were expanded post FACS, cells were plated in a 6 well format (220,000 cells per well). 24 hours after plating, cells were pelleted, and RNA extraction was immediately performed using the Qiagen RNeasy plus micro kit and QIAShredder kit. Using Applied Biosystems’ High-Capacity cDNA reverse transcription kit, cDNA synthesis was also performed on the same day as RNA extraction. PowerSYBR green PCR master mix from Applied Biosystems was used to perform standard comparative qPCR with primers for the two target and one housekeeping genes (*EGFR*, *TSPAN14*, and *GAPDH*). Three biological replicates in total were performed for each condition.

#### Extra information

Sequences of sgRNAs targeting rs7450443 w/o PAM sequence:

E1: TAGGCCTGAATGTCAATCAC
E2: AGTGTGTTGAGTGTGAACAC
E4: GTGTCAGCTCTCACTGAAAG

Sequences of non-targeting guides

N2: CGCTTCCGCGGCCCGTTCAA
NTC3: CCCGAGCAGTGGCTCGCTA

Sequence of positive control - sgRNA targeted to the enhancer of *TSPAN14* (Tspan14_enh): CTTAGGCGCTGCATACCGTA

## Data availability

The analyzed AD GWAS summary statistics data is available in GWAS catalog (https://ftp.ebi.ac.uk/pub/databases/gwas/summary_statistics/GCST90027001-GCST90028000/GCST90027158/). The selected FG datasets used in this study are available from the FILER database and in **Supplementary Table 2**.

## Code availability

Reported analyses were performed using the INFERNO pipeline (https://bitbucket.org/wanglab-upenn/bash-INFERNO).

## Authors’ contributions

YYL, PPK, AC, GS and L-SW conceived and designed the experiments. YYL, PPK, JC and LC performed data analyses. LC, JC, LB, SL, NT, SM, AR, KC, and RF carried out the data production and data generation under the supervision of YYL, SFAG, AC, FJ, GW, PDJ, JMV, LW. PPK, EG-A and OV provided IT support. YYL, PPK, SFAG, AC, and L-SW wrote and edited the manuscript. YYL, PDJ, JMV, SFAG, AC, GS and L-SW secured funding. All authors read and approved the manuscript.

## Acknowledgements

We would like to thank the members in the Alzheimer’s Disease Genetics Consortium for their feedback. The results published here are in whole or in part based on data obtained from Agora, a platform initially developed by the NIA-funded AMP-AD consortium that shares evidence in support of AD target discovery. Agora is available at: doi:10.57718/agora-adknowledgeportal.

