## Supplementary figures and tables for "Integrated genomic analysis and CRISPRi implicates *EGFR* in Alzheimer’s disease risk"

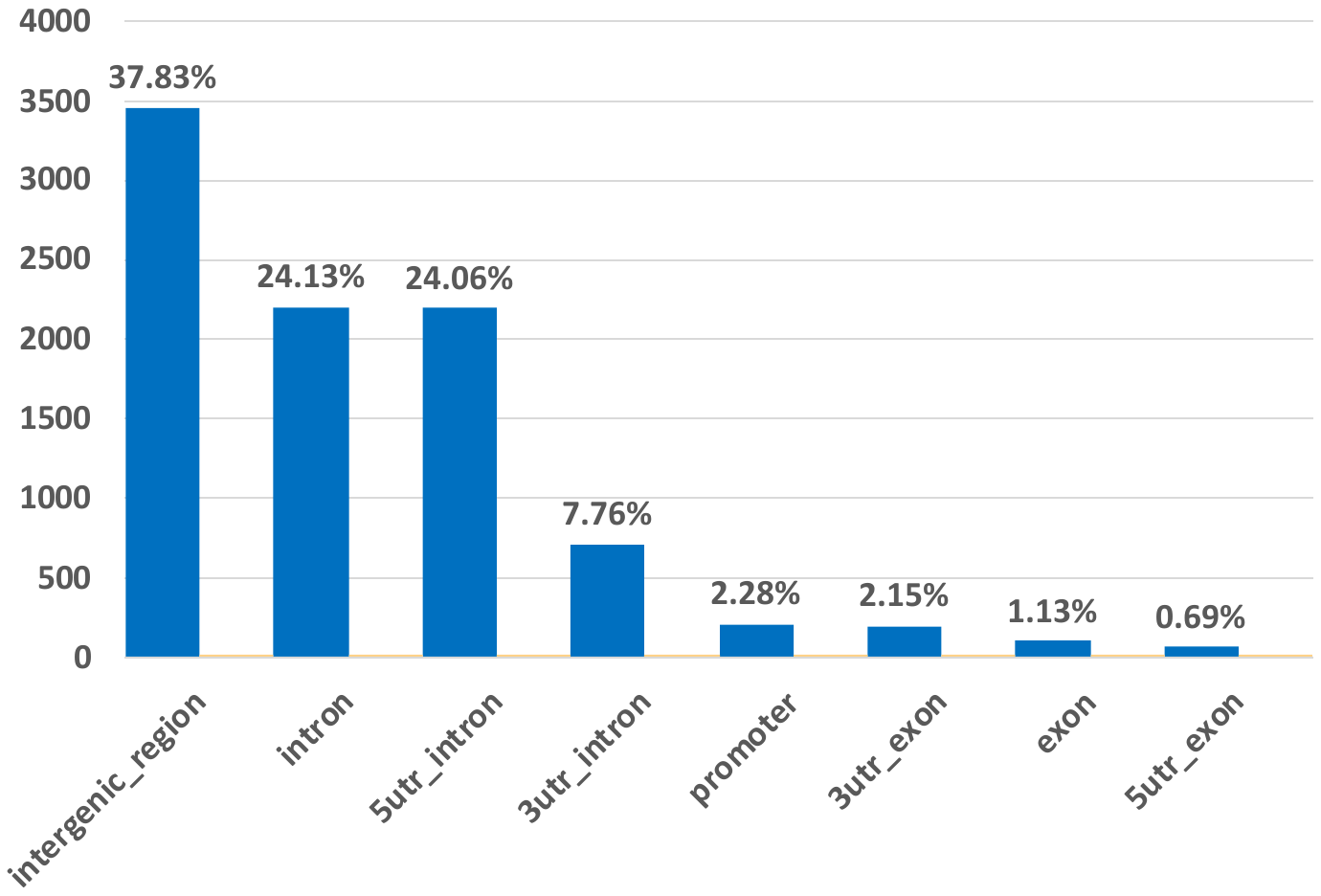

**Supplementary Fig. 1:** **Genomic localization of candidate regulatory variants**, shown as the proportion (%) in each category. Most (62%) are in intergenic and intronic regions, 35% in UTRs, and the rest in promoter and exonic regions.

A)

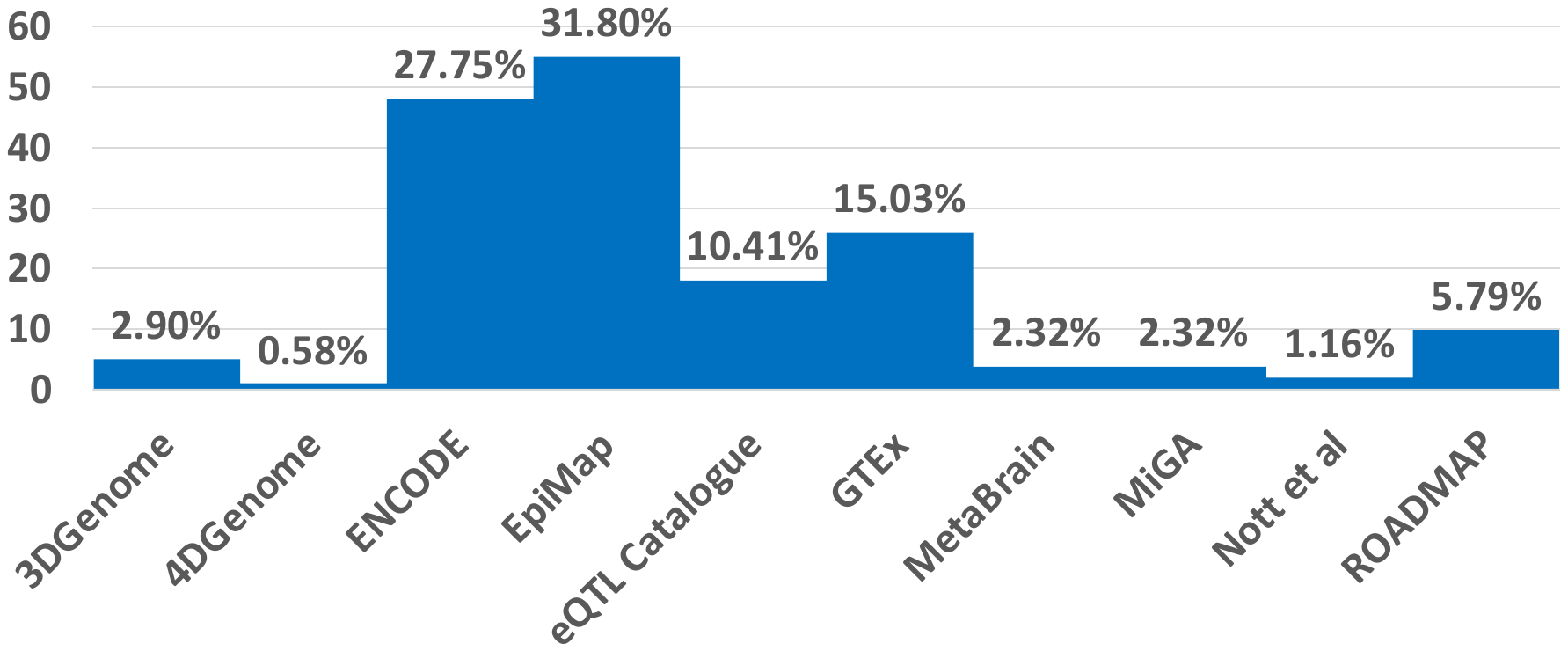

B)

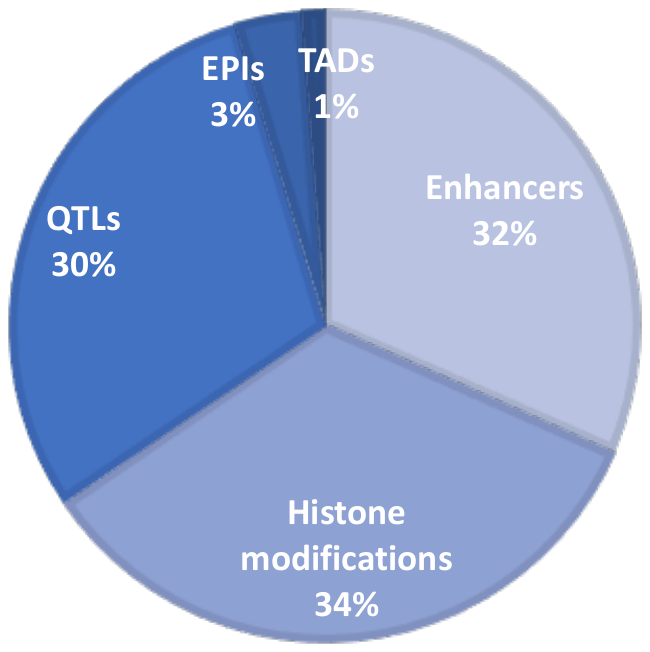

**Supplementary Fig. 2: Breadth of the FILER brain related tracks used for this analysis. A)** FILER brain related tracks by data sources. **B)** FILER brain related tracks by regulatory types.

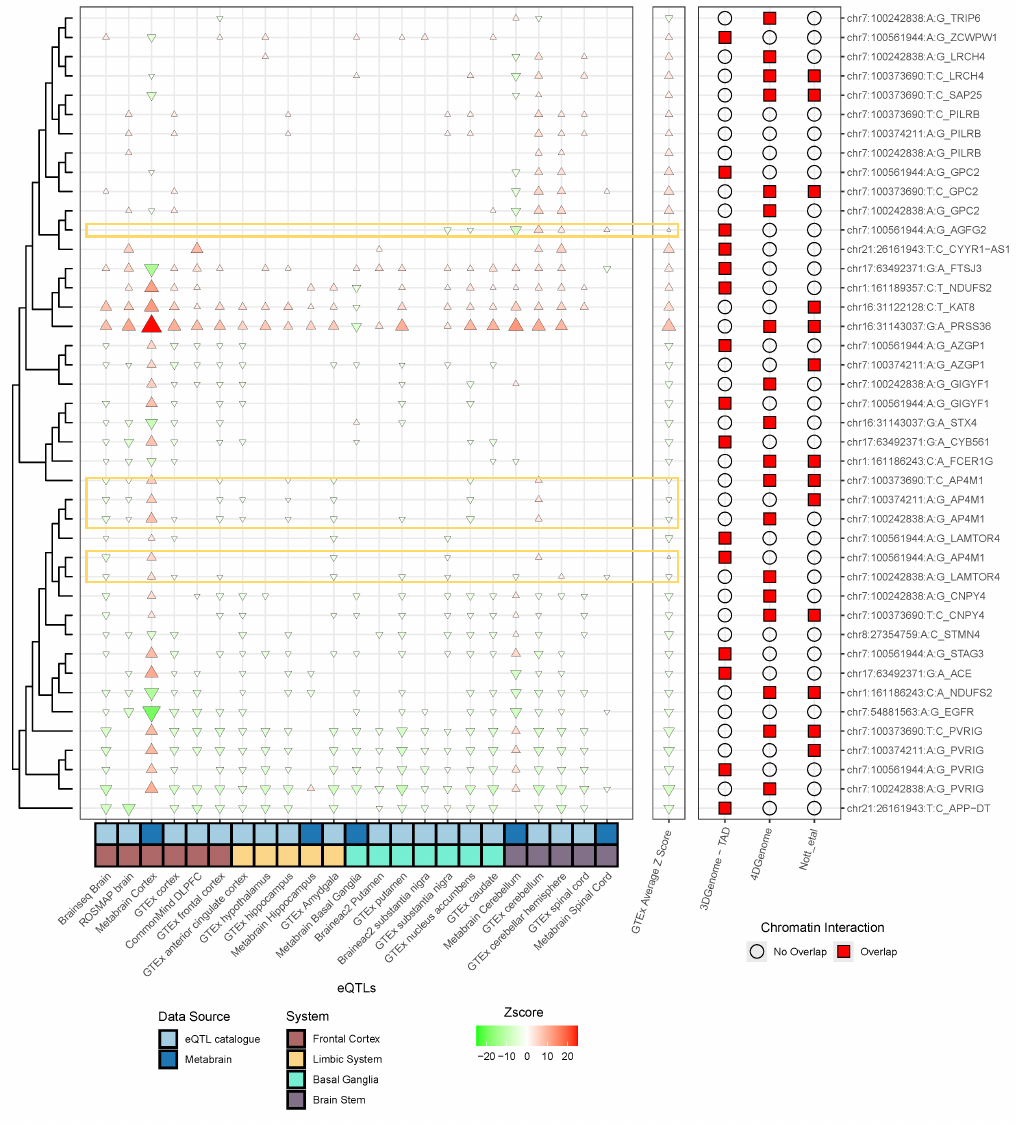

**Supplementary Fig. 3:** Comparison of directionality of selected putative V2G pairs across QTLs and EPI datasets used for *in silico* validation. Values in this plot are the original, non-normalized Z-scores (and therefore may be different from values in **Fig. 3**).

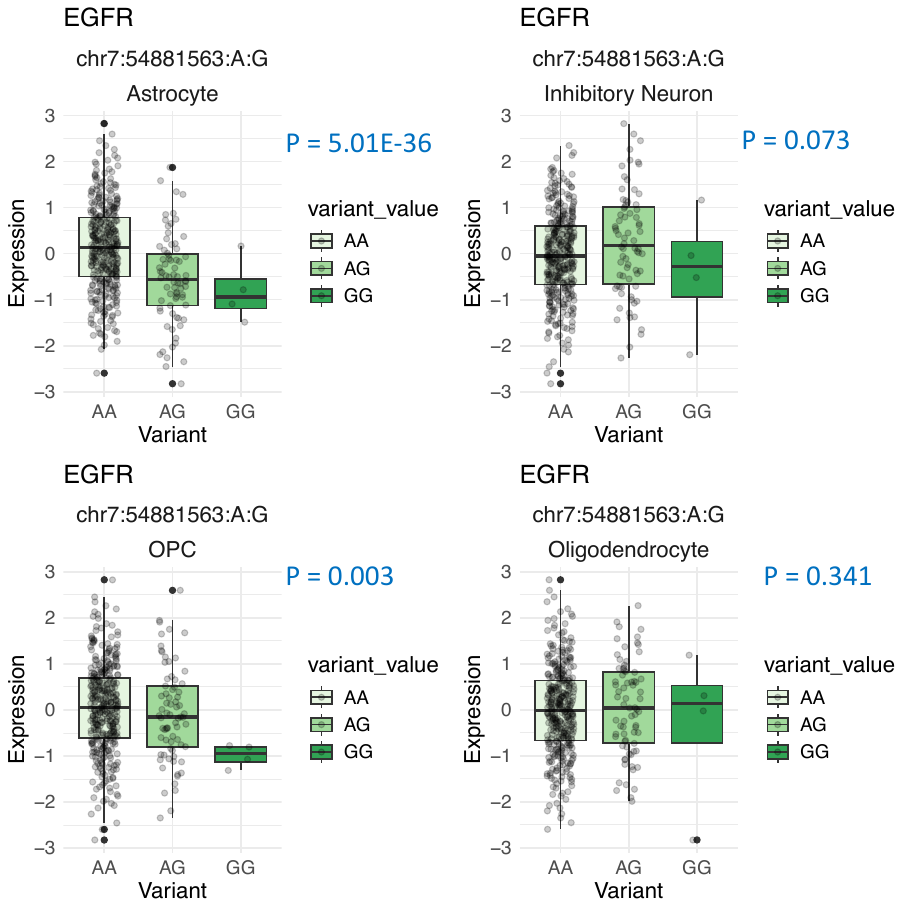

**Supplementary Fig. 4**. Single nuclei eQTL of rs74504435 and *EGFR* expression in four cell types (Astrocytes, Inhibitory Neurons, Oligodendrocyte precursor cells (OPCs), Oligodendrocytes) from ROSMAP.
